# Relative contribution of COVID-19 vaccination and SARS-CoV-2 infection to population-level seroprevalence of SARS-CoV-2 spike antibodies in a large integrated health system

**DOI:** 10.1101/2024.01.31.24301674

**Authors:** Tyler C. Chervo, Eric P. Elkin, Joshua R. Nugent, Emily Valice, Laura B. Amsden, Isaac J. Ergas, Julie R. Munneke, Monica Flores, Gina N. Saelee, Crystal A. Hsiao, Jeffery M. Schapiro, Charles P. Quesenberry, Douglas A. Corley, Laurel A. Habel, Lawrence H. Kushi, Jacek Skarbinski

**Affiliations:** Division of Research, Kaiser Permanente Northern California, Oakland, CA; The Permanente Medical Group, Kaiser Permanente Northern California, Oakland, CA; Department of Infectious Diseases, Oakland Medical Center, Kaiser Permanente Northern California, Oakland, CA; Physician Researcher Program, Kaiser Permanente Northern California, Oakland, CA

**Keywords:** SARS-CoV-2, seroprevalence, COVID-19 vaccination

## Abstract

**Background:** Understanding the relative contributions of SARS-CoV-2 infection-induced and vaccine-induced seroprevalence is key to measuring overall population-level seroprevalence and help guide policy decisions.

**Methods:** Using a series of six population-based cross-sectional surveys conducted among persons aged ≥7 years in a large health system with over 4.5 million members between May 2021 and April 2022, we combined data from the electronic health record (EHR), an electronic survey and SARS-CoV-2 spike antibody binding assay, to assess the relative contributions of infection and vaccination to population-level SARS-CoV-2 seroprevalence. EHR and survey data were incorporated to determine spike antibody positivity due to SARS-CoV-2 infection and COVID-19 vaccination. We used sampling and non-response weighting to create population-level estimates.

**Results:** We enrolled 4,319 persons over six recruitment waves. SARS-CoV-2 spike antibody seroprevalence increased from 83.3% (CI 77.0-88.9) in May 2021 to 93.5% (CI 89.5-97.5) in April 2022. By April 2022, 68.5% (CI 61.9-74.3) of the population was seropositive from COVID-19 vaccination only, 13.9% (10.7-17.9) from COVID-19 vaccination and prior diagnosed SARS-CoV-2 infection, 8.2% (CI 4.5- 14.5) from prior diagnosed SARS-CoV-2 infection only and 2.9% (CI 1.1-7.6) from prior undiagnosed SARS-CoV-2 infection only. We found high agreement (≥97%) between EHR and survey data for ascertaining COVID-19 vaccination and SARS-CoV-2 infection status.

**Conclusions:** By April 2022, 93.5% of persons had detectable SARS-CoV-2 spike antibody, predominantly from COVID-19 vaccination. In this highly vaccinated population and over 18 months into the pandemic, SARS-CoV-2 infection without COVID-19 vaccination was a small contributor to overall population-level seroprevalence.

**Article summary:** By April 2022, >93% of people had antibodies to SARS-CoV-2 with COVID-19 vaccination as the main driver of overall population-level seroprevalence in our healthcare system. SARS-CoV-2 infection without vaccination made a small contribution to population-level seroprevalence in our healthcare system.

## Background

By the end of January 2022, the United States has recorded over 75 million cases of coronavirus disease 2019 (COVID-19) and over 900,000 deaths.^1^ Antibodies that can neutralize severe acute respiratory syndrome coronavirus 2 (SARSl11CoVl112), the virus that causes COVID-19, can be generated either through SARS-CoV-2 infection or COVID-19 vaccination. A number of studies have reported high SARS-CoV-2 seroprevalence in the United States, but few studies have provided a detailed analysis of spike antibody seropositivity after vaccination and infection, differential seropositivity by demographic and clinical factors, or documented seropositivity over time.

In addition, seropositivity ascertainment may be biased by misclassification, and few studies have used multiple data sources (e.g., detailed electronic health records (EHR) and patient surveys) to comprehensively assess seropositivity in the population. Some studies have focused on easily accessible subsets of the population such as blood donors or commercial laboratory samples, which may not generalize well to other populations.^2–4^ In addition, despite numerous studies from the pre-vaccination era relatively few studies have been conducted when COVID-19 vaccines have been widely available.^5^

To address these gaps, we conducted six serial population-based cross-sectional serosurveys in a large integrated health system in California and combined data from the electronic health record (EHR) on COVID-19 vaccination and diagnosed SARS-CoV-2 infection, self-report of COVID-19 vaccination and SARS-CoV-2 infection on an electronic survey, and SARS-CoV-2 spike antibody binding assay test results to assess SARS-CoV-2 spike antibody seroprevalence over time. In addition, we examined the relative contributions of COVID-19 vaccination and SARS-CoV-2 infection to overall seroprevalence, identified demographic and clinical factors associated with seropositivity after COVID-19 vaccination or SARS-CoV- 2 infection, and assessed the agreement between EHR, survey and SARS-CoV-2 spike antibody testing to ascertain COVID-19 vaccination and SARS-CoV-2 infection status.

## Methods

### Setting

Kaiser Permanente Northern California (KPNC) is a large, integrated healthcare system that serves >4.5 million people in northern and central California. KPNC health plan members receive almost all preventive health services, including COVID-19 vaccination, and clinical care (e.g., SARS-CoV-2 diagnostic testing and treatment) within a closed network that includes 210 medical offices and 21 hospitals. KPNC provides health insurance and health care services to over 30% of the population within the counties in which KPNC has a physical presence. The demographic and socioeconomic characteristics of KPNC members closely approximate the region’s diverse demographics and includes patients with Medicare, Medicaid, and commercial insurance.^6^

### Study design, sampling, recruitment, and SARS-CoV-2 serological testing

We conducted six serial population-based cross-sectional studies that combine EHR clinical data, participant self-report from an electronic survey, and SARS-CoV-2 spike antibody testing to estimate SARS-CoV-2 spike antibody seroprevalence. For each wave of data collection, we developed a sampling frame of all KPNC members aged ≥7 years with active membership status for at least two months prior to the sampling date. We excluded persons who: 1) were known to be deceased on sampling date; 2) were known to have declined contact for all future studies at KPNC; 3) were previously sampled to participate in this study; 4) had no known email address on file; 5) were aged <18 years and did not have an identified parent or guardian in the EHR.

Using the sampling frame above, we selected six independent samples for six recruitment waves and sampling dates: Wave 1 (May 3^rd^ 2021), Wave 2 (May 25^th^ 2021), Wave 3 (July 29^th^ 2021), Wave 4 (September 8^th^ 2021), Wave 5 (October 12^th^ 2021), Wave 6 (December 20^th^ 2021). For each wave, we sampled 11,370 eligible persons using stratified random sampling with strata defined as age group in years (7-17, 18-44, 45-64, 65+) and race/ethnicity (white, Latinx/Hispanic, Asian, Black, other) combinations with selection probability proportional-to-size in all strata except Black persons. We oversampled Black persons in all age group strata three-fold to increase the sample size for this population. All activities involving human subjects were reviewed and ethically approved by the KPNC Institutional Review Board.

All sampled persons received up to three recruitment emails inviting them to participate in the survey. Persons who agreed to participate completed an electronic survey on demographics, COVID-19 prevention measures, self-reported vaccination status, and prior SARS-CoV-2 testing and infection. All persons who agreed to participate had a blood sample drawn for SARS-CoV-2 antibody testing at their local KPNC facility. Serology assays were conducted at our regional laboratory using the Diasorin Liaison SARS-CoV-2 S1/S2 IgG test which has been found to have a specificity of 98.9% and a sensitivity of 96.2%.^7,8^ This assay is used clinically in the KPNC health system, which provides a binary (positive/negative) result for the presence of SARS-CoV-2 spike binding antibody. Blood draws for each wave occurred during the following time periods: Wave 1 (May 2021 – August 2021; median May 5, 2021), Wave 2 (June 2021 – September 2021; median June 26, 2021), Wave 3 (August 2021 – February 2022; median August 27, 2021), Wave 4 (September 2021 – January 2022; median September 28, 2021), Wave 5 (October 2021 – February 2022; median October 30, 2021), and Wave 6 (January 2022 – April 2022; median January 24, 2022).

### Objectives and variable definitions

The primary objective was to estimate the prevalence of anti-SARS-CoV-2 spike binding antibody seropositivity and assess the relative contributions of COVID-19 vaccination, diagnosed SARS-CoV-2 infection, and undiagnosed SARS-COV-2 infection to overall population-level seroprevalence in our healthcare system. Diagnosed SARS-CoV-2 infection was based on having at least one of the following: a positive SARS-CoV-2 nucleic acid amplification test (NAAT) or antigen test in the EHR, a COVID-19 diagnosis in the EHR in absence of a positive lab result due to reporting a positive test, or self-report of COVID-19 in the survey. Undiagnosed SARS-CoV-2 infection was defined as a positive spike antibody test among participants with no COVID-19 vaccination or diagnosed SARS-CoV-2 infection. The secondary objectives were: 1) to assess factors associated with seropositivity among persons vaccinated for COVID- 19; 2) identify agreement between EHR and patient reported vaccination and infection status.

We assessed receipt of all doses of COVID-19 vaccines (BNT162b2 [Pfizer/BioNTech], mRNA- 1273 [Moderna/National Institutes of Health] or Ad.26.COV2.s [Janssen] prior to blood collection date in the EHR. Vaccination information was obtained from the EHR or from the California Immunization Registry; all vaccine providers in the state are required to report vaccine administration within 24 hours of administration. COVID-19 vaccination status was classified as: 1) Completed primary series, which includes persons who received one dose of Ad.26.COV2.s or 2 doses of BNT162b2 or m-RNA-1273; 2) Completed primary series plus an additional dose, which includes persons who completed a primary series and received at least one addition dose of any COVID-19 vaccine (Ad.26.COV2.s, BNT162b2 or m- RNA-1273); 3) Other, which includes persons who received one dose of either BNT162b2 or m-RNA- 1273, another vaccine product, or an unapproved combination of vaccines; 4) unvaccinated, which includes persons who have not received any COVID-19 vaccine. COVID-19 vaccination and SARS-CoV-2 infection were assessed via self-report in the survey using the question “Have you received a COVID-19 vaccine?” and “Have you ever been diagnosed with COVID-19?”, respectively.

Other variables of interest included demographic characteristics (age, sex, race/ethnicity), body mass index, Charlson comorbidity index score^9^ and individual Charlson comorbidities. Individual Charlson comorbidity categories were reclassified into the following: 1) atherosclerotic cardiovascular disease includes persons with prior myocardial infarction, congestive heart failure, peripheral vascular disease, and prior cerebrovascular event; 2) diabetes includes persons with diabetes with and without end-organ damage; 3) cancer includes persons with solid tumor with localized or metastatic disease, or leukemia and lymphoma.

### Weighting

Sampling weights were calculated as the inverse of the probability of selection within a given age and race strata. We adjusted for non-response bias by calculating a response weight defined as the inverse of the probability of having completed the survey and SARS-CoV-2 spike antibody testing. This probability was calculated using the “SuperLearner” package for the R programming language^10^, which evaluates several candidate algorithms for model fitting and creates a weighted ensemble model from these candidate algorithms. The variables used in non-response weighting were recruitment wave, age group, sex, race/ethnicity, geographic area, Charlson comorbidity index score category, preferred language, body mass index category, COVID-19 vaccination status and prior SARS-CoV2 infection in the EHR. Non-response weights were normalized to the size of the eligible sampled population and trimmed such that the maximum trimmed weight was the 99^th^ percentile of the untrimmed weights. The final weight used in the analysis was the product of the sampling weight and the non-response weight.

### Statistical methods

To estimate the prevalence of anti-SARS-CoV-2 spike antibody seropositivity in northern and central California, we calculated weighted frequencies and 95% confidence intervals (CIs). Contributions to population-level seroprevalence from prior infection, vaccination, or both were calculated as the percent of persons who tested positive for anti-SARS-CoV-2 spike antibodies and were in one of the above categories (ex. Antibody positive and has only been vaccinated). We assessed whether anti-SARS-CoV-2 seropositivity varied by levels of select demographic and clinical characteristics by performing a Chi-square test with Rao & Scott’s correction.^11^ To assess seropositivity among persons vaccinated for COVID-19, we used a modified log-Poisson regression with robust standard errors^12^ to estimate the adjusted prevalence ratio of anti-SARS-CoV-2 antibodies by age group, sex, race/ethnicity, Charlson comorbidity index score, chronic obstructive pulmonary disease, chronic kidney disease, atherosclerotic cardiovascular disease, cancer, diabetes, body mass index category, COVID-19 vaccination doses, product and months since last vaccine dose. Concordance between EHR and survey ascertainment of COVID-19 vaccination and SARS-CoV-2 infection was assessed by calculating unadjusted percentages. All analyses were performed in R version 4.0.2.

## Results

Of 3.5 million eligible KPNC members, we sampled 11,300 people per wave for a total of 67,800 people; of these sampled persons, 67,108 were found to be eligible during recruitment and 4,319 (6.4%) persons agreed to participate and completed the electronic survey and SARS-CoV-2 spike antibody testing (Supplementary Figure 1). Significant differences between respondents and non-respondents in each wave included recruitment wave, age group, sex, race/ethnicity, geographic area, Charlson comorbidity index score category, body mass index category, COVID-19 vaccination status and prior SARS-CoV2 infection in the EHR with standardized mean differences >0.20 (Supplementary Table 1). After applying non-response weights, the standardized mean differences between respondents and non-respondents across all characteristics were <0.20 except for combination of prior COVID-19 vaccination and SARS-CoV-2 infection at time of serology testing (0.28).

Across all six waves of data collection, 20% were aged 65+ years or older, 56% were female, 6.0% Black, 19% Hispanic, and 64% had a Charlson comorbidity index score of 0 (Table 1). In all, 75% had received COVID-19 vaccination only, 7.2% had received COVID-19 vaccination and had a diagnosed SARS-CoV-2 infection, 5.0% had prior diagnosed SARS-CoV-2 infection only and 13% had no COVID-19 vaccination or diagnosed SARS-CoV-2 infection. Overall, 69% had completed a primary COVID-19 vaccination series and 9.8% had completed a primary series and an additional dose; 45% received BNT162b2 (Pfizer/BioNTech) and 32% received mRNA-1273 (Moderna/NIH).

**Table 1.** Characteristics of eligible persons and prevalence of SARS-CoV-2 spike antibody positivity in in Kaiser Permanente Northern California, May 2021 – April 2022.

| Characteristic | Population |  | SARS-CoV-2 spike antibody positive |  |  |
| --- | --- | --- | --- | --- | --- |
|  | N | Weighted population size (column %) | N | Weighted % (CI) | p-value |
| **Total** | 4,319 | 3,589,422 (100%) | 4,104 | 87.5% (85.5, 89.4) |  |
| **Recruitment wave** |  |  |  |  | 0.020 |
| 1 (May 2021 - August 2021) | 458 | 3,074,884 (14%) | 420 | 83.3% (77.9, 88.8) |  |
| 2 (June 2021 - September 2021) | 556 | 3,530,422 (16%) | 518 | 82.9% (77, 88.9) |  |
| 3 (August 2021 - February 2022) | 940 | 3,770,016 (18%) | 885 | 86% (81.8, 90.1) |  |
| 4 (September 2021 - January 2022) | 897 | 3,752,643 (18%) | 854 | 88.1% (84.1, 92.2) |  |
| 5 (October 2021 - February 2022) | 717 | 3,817,245 (18%) | 689 | 90.3% (85.8, 94.7) |  |
| 6 (January 2022 - April 2022) | 751 | 3,482,437 (16%) | 738 | 93.5% (89.5, 97.5) |  |
| **Age group (years)** |  |  |  |  | <0.001 |
| 7-17 | 72 | 253,384 (7.0%) | 48 | 63.3% (50.2, 76.4) |  |
| 18-44 | 1,062 | 1,475,697 (41%) | 986 | 86.3% (83, 89.6) |  |
| 45-64 | 1,743 | 1,160,236 (32%) | 1,664 | 90.1% (87.7, 92.6) |  |
| 65+ | 1,442 | 700,106 (20%) | 1,406 | 94.1% (91.6, 96.7) |  |
| **Sex** |  |  |  |  | >0.9 |
| Female | 2,751 | 1,988,539 (56%) | 2,622 | 87.4% (84.9, 89.9) |  |
| Male | 1,568 | 1,600,883 (44%) | 1,482 | 87.5% (84.6, 90.5) |  |
| **Race/Ethnicity** |  |  |  |  | 0.011 |
| White | 2,947 | 1,631,062 (45%) | 2,794 | 85.3% (82.9, 87.7) |  |
| Asian | 493 | 755,468 (21%) | 484 | 94.3% (90.4, 98.1) |  |
| Black | 352 | 215,929 (6.0%) | 325 | 80.6% (74.1, 87.2) |  |
| Hispanic | 371 | 665,137 (19%) | 356 | 89.4% (84.2, 94.6) |  |
| Other | 156 | 321,826 (8.9%) | 145 | 82.9% (73.1, 92.7) |  |
| **Geographic area** |  |  |  |  | 0.003 |
| Central valley | 326 | 357,636 (9.9%) | 298 | 81.1% (73.4, 88.8) |  |
| East bay | 1,169 | 1,004,297 (28%) | 1,129 | 90.9% (87.7, 94.1) |  |
| North bay | 458 | 244,449 (6.9%) | 442 | 92% (87.9, 96.1) |  |
| North valley | 1,220 | 978,828 (27%) | 1,136 | 83.7% (79.9, 87.5) |  |
| Peninsula | 516 | 411,390 (12%) | 502 | 93.5% (89.2, 97.7) |  |
| South bay | 630 | 592,821 (16%) | 597 | 85.6% (79.9, 91.2) |  |
| **Charlson score category** |  |  |  |  | 0.3 |
| No visits in prior year | 125 | 280,587 (7.8%) | 114 | 83.9% (74.2, 93.6) |  |
| Score 0 | 2,647 | 2,315,506 (64%) | 2,510 | 86.6% (84.2, 89.1) |  |
| Score 1 | 749 | 482,293 (13%) | 720 | 89.9% (85.8, 94) |  |
| Score 2 | 370 | 198,729 (5.6%) | 355 | 91.6% (86.9, 96.3) |  |
| Score 3+ | 428 | 312,307 (8.8%) | 405 | 90.4% (85.1, 95.6) |  |
| **Chronic obstructive pulmonary disease** | 582 | 388,464 (11%) | 557 | 90.4% (86.1, 94.8) | 0.2 |
| **Chronic kidney disease** | 246 | 173,981 (4.9%) | 233 | 88% (79.6, 96.4) | 0.9 |
| **Atherosclerotic cardiovascular disease** | 769 | 425,534 (12%) | 736 | 91.3% (88.2, 94.5) | 0.039 |
| **Cancer/Malignancies** | 196 | 117,166 (3.3%) | 184 | 93% (88.8, 97.1) | 0.047 |
| **Diabetes** | 441 | 342,493 (9.6%) | 425 | 91.7% (86.6, 96.7) | 0.2 |
| **BMI category** |  |  |  |  | <0.001 |
| 18.5 - <25 | 1,281 | 966,367 (27%) | 1,228 | 90% (86.8, 93.2) |  |
| 25 - <30 | 1,476 | 1,088,391 (30%) | 1,416 | 91.2% (88.6, 93.8) |  |
| 30 - <40 | 1,174 | 951,293 (27%) | 1,105 | 83.9% (79.9, 88) |  |
| >= 40 | 254 | 189,514 (5.3%) | 243 | 91.4% (85.7, 97.1) |  |
| Unknown | 134 | 393,857 (11%) | 112 | 77.4% (68.2, 86.7) |  |
| **COVID-19 vaccination or diagnosed SARS-CoV-2 infection prior to spike antibody testing (EHR and survey)** |  |  |  |  | <0.001 |
| Neither | 195 | 460,642 (13%) | 22 | 12.7% (6.7, 18.7) |  |
| Prior infection only | 81 | 178,615 (5.0%) | 70 | 88.5% (81, 96) |  |
| Vaccinated and prior infection | 326 | 257,045 (7.2%) | 326 | 100% (100, 100) |  |
| Vaccinated only | 3,717 | 2,693,119 (75%) | 3,686 | 99% (98.5, 99.5) |  |
| **Vaccination series** |  |  |  |  | <0.001 |
| Not vaccinated | 289 | 669,857 (19%) | 103 | 36.3% (29.3, 43.4) |  |
| Partial | 43 | 95,977 (2.8%) | 42 | 97% (91.2, 100) |  |
| Primary series complete | 3,162 | 2,477,006 (69%) | 3,135 | 99.2% (98.8, 99.6) |  |
| Primary series + additional dose | 825 | 346,583 (9.8%) | 824 | 99.9% (99.7, 100) |  |
| **Vaccination status at blood draw time** |  |  |  |  | <0.001 |
| Ad26.COV2.S (Janssen) | 192 | 148,530 (4.2%) | 187 | 96.1% (91.2, 100) |  |
| BNT162b2 (Pfizer/BioNTech) | 2,107 | 1,619,133 (45%) | 2,094 | 99.5% (99.2, 99.8) |  |
| mRNA-1273 (Moderna/NIH) | 1,731 | 1,151,903 (32%) | 1,720 | 99.2% (98.6, 99.8) |  |
| Unvaccinated | 289 | 669,857 (19%) | 103 | 36.3% (29.3, 43.4) |  |
| **Vaccination in medical records** | 4,030 | 2,919,565 (81%) | 4,001 | 99.2% (98.8, 99.6) | <0.001 |
| **COVID-19 diagnosis in medical records** | 311 | 303,276 (8.5%) | 303 | 95.3% (91.7, 99) | 0.005 |
| **Indicated vaccination in survey** | 3,927 | 2,861,173 (80%) | 3,900 | 99.3% (98.9, 99.6) | <0.001 |
| **Indicated infection in survey** | 375 | 405,050 (11%) | 365 | 95.8% (92.9, 98.7) | <0.001 |

CI=95% confidence interval; EHR=electronic health record
P-values calculated using chi-squared test with Rao & Scott's correction for assessing the distribution of SARS-CoV-2 spike antibody positivity by population characteristics.

Across all six waves, 87.5% had detectable SARS-CoV-2 spike antibody, with a range of 83.3% in Wave 1 (median test date May 5, 2021) to 93.5% in Wave 6 (median test date January 24, 2022). We found differences in seroprevalence by age group (e.g., aged 7-17 years 63.3% versus aged 65+ years 94.1%) and race/ethnicity (Black 80.6% versus Asian 94.3%). Seroprevalence was higher among persons who received COVID-19 vaccination only (99.0%) versus persons with prior diagnosed SARS-COV-2 infection only (88.5%). Among persons who had no prior COVID-19 vaccination or diagnosed SARS-CoV-2 infection, 12.7% had detectable SARS-CoV-2 spike antibody.

Within our healthcare system, from May 2021 to April 2022, SARS-CoV-2 infection incidence peaked in August 2021 (during circulation of the Delta variant; B.1.617.2) and January 2022 (during circulation of the Omicron variant; B.1.1.529) (Figure 1, panel A) and the percentage of all persons in our health system who received at least one COVID-19 vaccination increased from 38% in May 2021 to 70.4% in February 2022 (Figure 1, panel B). COVID-19 vaccination was the main driver of increasing seroprevalence in our healthcare system (Table 2). The contribution to population level seroprevalence by people with diagnosed SARS-CoV-2 infection increased from 8.3% in Wave 1 to 22.1% by Wave 6. Overall, only a small percentage had undiagnosed SARS-CoV-2 infection (positive spike antibody test among participants with no COVID-19 vaccination or diagnosed SARS-CoV-2 infection) (e.g., 2.9% in wave 6).

**Figure 1.**
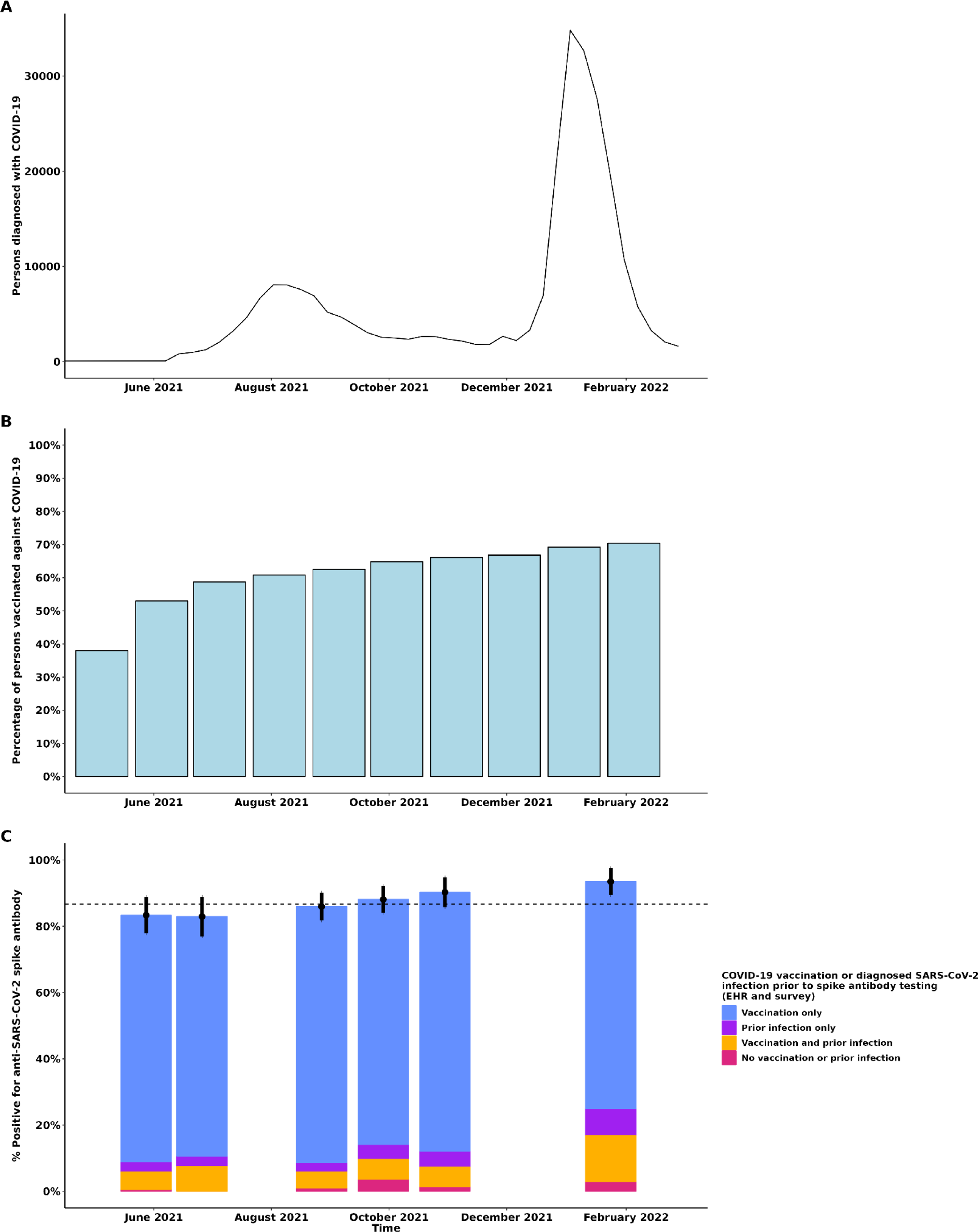
Panel A. COVID-19 cases diagnosed in Kaiser Permanente Northern California June 2021 – February 2022 by week. **Panel B.** Percentage of Kaiser Permanente Northern California members who have received at least one COVID-19 vaccine May 2021 – February 2022. **Panel C.** Relative contributions of COVID-19 vaccination, and diagnosed and undiagnosed SARS-CoV-2 infection to population-level prevalence of anti-SARS-CoV-2 spike protein antibody by recruitment wave, May 2021 – April 2022. The dashed line in Panel C represents the average level of seroprevalence across the study’s duration. Note: COVID-19 vaccination based on electronic health record or self-report on survey. Diagnosed SARS-CoV-2 infection was defined as a positive SARS-CoV-2 nucleic acid amplification test (NAAT) or antigen test recorded in the EHR or self- report of COVID-19 in the survey. Undiagnosed SARS-CoV-2 infection was defined as a positive spike antibody test among participants with no COVID-19 vaccination or diagnosed SARS-CoV-2 infection. COVID-19 vaccination status was classified as: 1) Completed primary series, which includes persons who received one dose of Ad.26.COV2.s or 2 doses of BNT162b2 or m-RNA-1273; 2) Completed primary series plus an additional dose, which includes persons who completed a primary series and received at least one addition dose of any COVID-19 vaccine (Ad.26.COV2.s, BNT162b2 or m-RNA-1273); 3) Other, which includes persons who received one dose of either BNT162b2 or m-RNA-1273, another vaccine product, or an unapproved combination of vaccines.

**Table 2.**
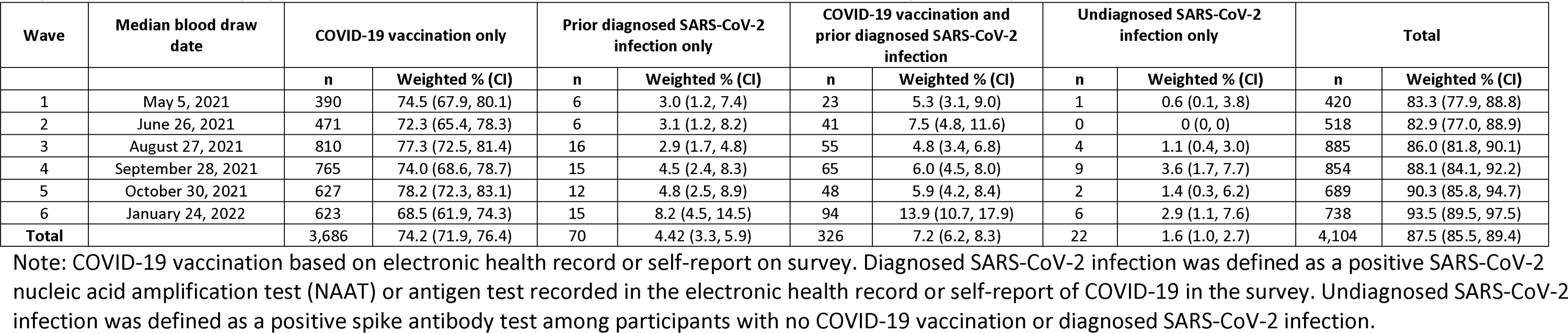
Relative contributions of COVID-19 vaccination, and diagnosed and undiagnosed SARS-CoV-2 infection to population-level prevalence of anti-SARS-CoV- 2 spike protein antibody by recruitment wave in Kaiser Permanente Northern California, May 2021 – April 2022.

| Wave | Median blood draw date | COVID-19 vaccination only |  | Prior diagnosed SARS-CoV-2 infection only |  | COVID-19 vaccination and prior diagnosed SARS-CoV-2 infection |  | Undiagnosed SARS-CoV-2 infection only |  | Total |  |
| --- | --- | --- | --- | --- | --- | --- | --- | --- | --- | --- | --- |
|  |  | n | Weighted % (CI) | n | Weighted % (CI) | n | Weighted % (CI) | n | Weighted % (CI) | n | Weighted % (CI) |
| 1 | May 5, 2021 | 390 | 74.5 (67.9, 80.1) | 6 | 3.0 (1.2, 7.4) | 23 | 5.3 (3.1, 9.0) | 1 | 0.6 (0.1, 3.8) | 420 | 83.3 (77.9, 88.8) |
| 2 | June 26, 2021 | 471 | 72.3 (65.4, 78.3) | 6 | 3.1 (1.2, 8.2) | 41 | 7.5 (4.8, 11.6) | 0 | 0 (0, 0) | 518 | 82.9 (77.0, 88.9) |
| 3 | August 27, 2021 | 810 | 77.3 (72.5, 81.4) | 16 | 2.9 (1.7, 4.8) | 55 | 4.8 (3.4, 6.8) | 4 | 1.1 (0.4, 3.0) | 885 | 86.0 (81.8, 90.1) |
| 4 | September 28, 2021 | 765 | 74.0 (68.6, 78.7) | 15 | 4.5 (2.4, 8.3) | 65 | 6.0 (4.5, 8.0) | 9 | 3.6 (1.7, 7.7) | 854 | 88.1 (84.1, 92.2) |
| 5 | October 30, 2021 | 627 | 78.2 (72.3, 83.1) | 12 | 4.8 (2.5, 8.9) | 48 | 5.9 (4.2, 8.4) | 2 | 1.4 (0.3, 6.2) | 689 | 90.3 (85.8, 94.7) |
| 6 | January 24, 2022 | 623 | 68.5 (61.9, 74.3) | 15 | 8.2 (4.5, 14.5) | 94 | 13.9 (10.7, 17.9) | 6 | 2.9 (1.1, 7.6) | 738 | 93.5 (89.5, 97.5) |
| **Total** |  | 3,686 | 74.2 (71.9, 76.4) | 70 | 4.42 (3.3, 5.9) | 326 | 7.2 (6.2, 8.3) | 22 | 1.6 (1.0, 2.7) | 4,104 | 87.5 (85.5, 89.4) |

Note: COVID-19 vaccination based on electronic health record or self-report on survey. Diagnosed SARS-CoV-2 infection was defined as a positive SARS-CoV-2 nucleic acid amplification test (NAAT) or antigen test recorded in the electronic health record or self-report of COVID-19 in the survey. Undiagnosed SARS-CoV-2 infection was defined as a positive spike antibody test among participants with no COVID-19 vaccination or diagnosed SARS-CoV-2 infection.

We assessed factors associated with detectable SARS-CoV-2 spike antibodies among persons who received a complete primary COVID-19 vaccination series with or without an additional dose and who had no prior diagnosed SARS-COV-2 infection (Table 3). Overall, 99% had detectable SARS-CoV-2 spike antibodies. Factors significantly associated with detectable SARS-CoV-2 spike antibodies among vaccinated persons were Asian race (adjusted prevalence ratio [aPR] 1.01; CI 1.00-1.01) and Black race (aPR 1.01; CI 1.01-1.02) relative to White race and having completed a primary COVID-19 vaccination series plus an additional dose relative to having completed a primary series only (aPR 1.01; CI 1.00-1.02).

**Table 3.** SARS-CoV-2 spike antibody seropositivity and factors associated with seropositivity among persons with SARS-CoV-2 spike protein seropositivity among persons who completed at least a COVID-19 primary vaccination series and had no history of diagnosed SARS-CoV-2 infection, Kaiser Permanente Northern California, May 2021 – January 2022

| Characteristic | N =<br>3,739 | % Seropositive (95% CI) | Prevalence Ratio | 95% CI | p-value |
| --- | --- | --- | --- | --- | --- |
| **Age group (years)** |  |  |  |  |  |
| 18-44 | 816 | 98.5% (95.4, 100) | — | — |  |
| 7-17 | 33 | 99.5% (98.8, 100) | 0.99 | 0.96, 1.02 | 0.38 |
| 45-64 | 1,476 | 99.3% (98.9, 99.8) | 1.00 | 0.99, 1.01 | 0.73 |
| 65+ | 1,344 | 98.7% (97.9, 99.4) | 1.01 | 1.00, 1.02 | 0.11 |
| **Sex** |  |  |  |  |  |
| Male | 1,354 | 99.5% (99.2, 99.8) | — | — |  |
| Female | 2,315 | 98.8% (98.1, 99.6) | 1.01 | 1.00, 1.01 | 0.24 |
| **Race/Ethnicity** |  |  |  |  |  |
| White | 2,544 | 98.7% (98.2, 99.3) | — | — |  |
| Asian | 445 | 100% (100, 100) | 1.01 | 1.00, 1.01 | 0.002 |
| Black | 274 | 100% (100, 100) | 1.01 | 1.01, 1.02 | <0.001 |
| Hispanic | 277 | 99.1% (97.3, 100) | 1.00 | 0.99, 1.02 | 0.81 |
| Other | 129 | 99.3% (97.9, 100) | 1.00 | 0.98, 1.02 | 0.77 |
| **Charlson score category** |  |  |  |  |  |
| Score 0 | 2,205 | 100% (100, 100) | — | — |  |
| Score 1 | 651 | 99.5% (99, 100) | 1.00 | 0.98, 1.01 | 0.79 |
| Score 2 | 330 | 99.8% (99.5, 100) | 1.00 | 0.97, 1.04 | 0.86 |
| Score 3+ | 379 | 99.2% (98.4, 100) | 0.99 | 0.94, 1.05 | 0.75 |
| No visits in prior year | 104 | 95.8% (93.5, 98.1) | 1.01 | 1.00, 1.01 | 0.050 |
| **Chronic obstructive pulmonary disease** | 495 | 99.2% (98.4, 99.9) | 1.01 | 0.99, 1.02 | 0.41 |
| **Chronic kidney Disease** | 225 | 95.3% (91.9, 98.7) | 0.97 | 0.92, 1.01 | 0.17 |
| **Atherosclerotic cardiovascular disease** | 686 | 97.1% (95.5, 98.7) | 0.98 | 0.97, 1.00 | 0.11 |
| **Cancer/Malignancies** | 179 | 92.7% (88.3, 97.1) | 0.94 | 0.89, 1.00 | 0.066 |
| **Diabetes** | 379 | 99.3% (98.5, 100) | 1.02 | 0.99, 1.04 | 0.25 |
| **BMI Category** |  |  |  |  |  |
| 18.5 - <25 | 1,121 | 99.3% (98.9, 99.8) | — | — |  |
| 25 - <30 | 1,282 | 99.4% (99, 99.8) | 1.00 | 1.00, 1.01 | 0.29 |
| 30 - <40 | 970 | 98.8% (97.6, 100) | 1.00 | 0.99, 1.01 | 0.89 |
| >= 40 | 202 | 99.1% (98.1, 100) | 1.00 | 0.98, 1.01 | 0.59 |
| Unknown | 94 | 99.4% (98.2, 100) | 1.00 | 1.0, 1.01 | 0.41 |
| **Vaccine type at enrollment** |  |  |  |  |  |
| BNT162b2 (Pfizer/BioNTech) | 1,910 | 95.7% (90.3, 100) | — | — |  |
| Ad26.COV2.S (Janssen) | 163 | 99.4% (99.1, 99.8) | 0.96 | 0.91, 1.02 | 0.18 |
| mRNA-1273 (Moderna/NIH) | 1,596 | 99.4% (99, 99.8) | 1.00 | 0.99, 1.01 | 0.83 |
| **Vaccination status at blood draw time** |  |  |  |  |  |
| Primary series complete | 2,909 | 99.1% (98.7, 99.6) | — | — |  |
| Primary series + additional dose | 760 | 99.9% (99.7, 100) | 1.01 | 1.00, 1.02 | 0.019 |
| **Months since last vaccination** |  |  |  |  |  |
| <1 month | 334 | 100% (100, 100) | — | — |  |
| 1 - 2 months | 940 | 99.2% (98.6, 99.8) | 1.00 | 0.99, 1.01 | 0.80 |
| 3 - 4 months | 974 | 98.8% (97.7, 99.9) | 0.99 | 0.99, 1.00 | 0.17 |
| >=5 months | 1,421 | 99.4% (98.9, 99.9) | 1.00 | 0.99, 1.01 | 0.80 |

CI = 95% confidence interval; EHR = electronic health record
Model adjusting for age group in years, sex, race/ethnicity, charlson comorbidity index score, chronic obstructive pulmonary disease status, chronic kidney disease status, atherosclerotic cardiovascular disease status, cancer status, diabetes status, body mass index category, COVID-19 vaccination series in the EHR, COVID-19 vaccination product in the EHR, and months since vaccination

For ascertainment of COVID-19 vaccination status and prior diagnosed SARS-CoV-2 infection, the EHR and patient self-report had 99.1% agreement for COVID-19 vaccination and 97.1% for prior infection (Table 4). However, the EHR only identified 74.4% of diagnosed prior infections (sensitivity) compared to the survey using self-report as the gold standard. For ascertainment of prior SARS-CoV-2 spike antigen exposure as measured by SARS-CoV-2 spike antibody testing, spike antibody test results had 98.5% agreement with EHR or patient self-report of COVID-19 vaccination or prior diagnosed infection (Table 5).

**Table 4.** Concordance of data from electronic health record and patient self-report on electronic survey for COVID-19 vaccination and prior diagnosed SARS-CoV-2 infection status in Kaiser Permanente Northern California, May 2021 – April 2022 (N=4,220)

|  | **COVID-19**<br>**vaccination** | **Prior diagnosed SARS-**<br>**CoV-2 infection** |
| --- | --- | --- |
| **In EHR and survey (Concordant)** | 3914 | 279 |
| **Not in EHR and not in survey (Concordant)** | 271 | 3818 |
| **In EHR and not in survey (Discordant)** | 22 | 27 |
| **Not in EHR, but in survey (Discordant)** | 13 | 96 |
| **Sensitivity** | 99.7% | 74.4% |
| **Specificity** | 92.5% | 99.3% |
| **Positive predictive value** | 99.4% | 88.2% |
| **Negative predictive value** | 95.6% | 98.2% |
| **Agreement** | 99.1% | 97.1% |

EHR = electronic health record
Note: Sensitivity, specificity, positive and negative predictive value calculated using self-reported data from electronic survey on COVID-19 vaccination and prior diagnosed SARS-CoV-2 infection as the “gold standard” Agreement is defined as sum of concordant results over the total.

**Table 5:** Concordance of COVID-19 vaccination and diagnosed SARS-CoV-2 infection status and SARS-CoV-2 spike antibody results in Kaiser Permanente Northern California, May 2021 – April 2022 (N=4,319)

|  | **Spike antibody positive** |
| --- | --- |
| **Prior COVID-19 vaccination or SARS-CoV-2 infection and spike antibody positive (Concordant)** | 4082 |
| **No prior COVID-19 vaccination or SARS-CoV-2 infection and spike antibody negative (Concordant)** | 173 |
| **Prior COVID-19 vaccination or SARS-CoV-2 infection and spike antibody negative (Discordant)** | 42 |
| **No prior COVID-19 vaccination or SARS-CoV-2 infection and spike antibody positive (Discordant)** | 22 |
| **Sensitivity** | 99.5% |
| **Specificity** | 80.5% |
| **Positive predictive value** | 98.9% |
| **Negative predictive value** | 89.7% |
| **Agreement** | 98.5% |

EHR = electronic health record
Note: Sensitivity, specificity, positive and negative predictive value calculated using SARS-CoV-2 spike antibody test results as the “gold standard”
Agreement is defined as sum of concordant results divided by the total.

## Discussion

In this series of six cross-sectional population-based serosurveys with linked EHR and self-reported survey data in a large integrated health system in California, we found that by the end of our sixth recruitment wave in April 2022, 93.5% of persons were seropositive for spike antibody, with prior COVID-19 vaccination only being the main contributor (68.5%), followed by diagnosed SARS-COV-2 COVID-19 vaccination and diagnosed SARS-CoV-2 infection (13.9%) and diagnosed SARS-CoV-2 infection only (8.2%); undiagnosed SARS-CoV-2 infection among unvaccinated persons contributed a small proportion to population level seroprevalence (2.9%). Among persons without prior diagnosed infection who have received at least a primary COVID-19 vaccination series, 99% had detectable SARS-COV-2 antibodies with minimal differences among sub-groups (e.g., 99.2% among person who received primary series only and 99.9% among person who received a primary series plus an additional dose).

Numerous seroprevalence studies have been conducted to date.^2,13–16^ Similar to this study, others^14,16^ conducted in California around the time of our study period have found high levels of seroprevalence with >90% of persons seropositive for spike antibody by early 2022 due to exposure to COVID-19 vaccination or SARS-CoV-2 infection. Similarly, to other studies, we found that vaccination was the main driver of population-level seroprevalence, but that by April 2022 a substantial percentage of the population had a history of SARS-COV-2 infection (25%). Although, we did not conduct testing for SARS-CoV-2 infection with nucleocapsid antibody testing, we were able to identify a small percentage of persons who have undiagnosed SARS-CoV-2 infection among those with no recorded or self-reported SARS-CoV-2 infection or vaccination. The estimated fraction of undiagnosed infections has varied across studies, with one US population-based study from early in the pandemic finding an undiagnosed seropositivity rate of 4.6% by mid-July 2020.^17^ Another study conducted in California June – August 2020 estimated that only 31% of infections had been reported to the California Department of Public Health.^13^ By the end of our study period, we found that the overall weighted seropositivity among persons without EHR confirmed or self-reported COVID-19 vaccination or SARS-CoV-2 infection to be 12.6%, though we note that we could not identify the prevalence of undiagnosed infections among vaccinated persons. The combined changes in the availability of COVID-19 testing over time and increasing transmissibility of SARS-CoV-2 variants will change the proportion of infections that go undiagnosed over time and further studies of this type, including among vaccinated persons, are important to estimating this proportion.

Seroprevalence was high among most demographic groups, except for children aged 7-17 years; this likely represents lower COVID-19 vaccination rates as recommendations for COVID-19 vaccination lagged for children compared to adults. In addition, seropositivity was lower among Black Americans (80.6%) compared with Asian Americans (94.3%) suggesting racial/ethnicity disparities in COVID-19 vaccination rates since vaccination is the major driver of seroprevalence in our healthcare system. Other research has shown disparities in COVID-19 vaccine uptake by race due to a variety of causes.^18,19^ Addressing disparities in vaccine uptake among different race/ethnic groups will contribute to reducing disparities in COVID-19 morbidity and mortality.^20^

Seropositivity among persons who received COVID-19 vaccination was very high (99%) overall and minimal differences were noted by demographic or clinical characteristics. The commercially available assay used in this study is highly sensitive and seropositivity after vaccination is almost uniformly high. However, these binding antibody assays might not correlate with viral neutralization assays or accurately predict protection from future SARS-CoV-2 infection or severe COVID-19 and thus have limited clinical utility for assessing vaccine effectiveness.

Lastly, we found high levels of agreement between EHR and patient self-report via survey to ascertain prior COVID-19 vaccination or diagnosed SARS-CoV-2 infection as well as agreement between EHR or self-reported prior SARS-CoV2 spike antigen exposure and spike antibody test results. Given the transition in COVID-19 surveillance with ever greater reliance on EHR data to monitor COVID-19 vaccination or SARS-COV-2 infections to guide policy decisions, this study provides reassurance that EHRs can be used for SARS-CoV-2 disease monitoring. These findings are comparable to other studies that have evaluated the concordance of EHR and survey data for SARS-CoV-2 infection.^21^

This study has several limitations. First, we only used spike antibody testing, which can be positive after either COVID-19 vaccination or SARS-CoV-2 infection but did not use an antibody test targeting the nucleocapsid antigen that can be used to detect SARS-CoV-2 infection alone. This has limited our ability to detect the full set of undiagnosed SARS-CoV-2 infections, particularly among vaccinated persons, and thus our estimate of persons with undiagnosed SARS-CoV-2 infection across the duration of the study is likely an underestimate. Second, although we have used standard population-based sampling and weighting methods, our overall response rate is modest and thus the respondent population may not be representative of the general population. This was mitigated in part by use of detailed EHR data on all sampled persons to create non-response weights. Additionally, some patients experience with SARS-CoV-2 may have made them more likely to remember an infection than others, which could potentially bias the results of the study.

In conclusion, we found that by the end of our study, 93.5% of persons were seropositive for spike antibody with the majority of seropositivity coming from COVID-19 vaccination. Seroprevalence was high among all groups except children aged 7-17 years, and almost all persons had detectable SARS- CoV-2 spike antibodies after COVID-19 vaccination. Within our healthcare system, COVID-19 vaccination was the main contributor to overall population-level seroprevalence.

## Data Availability

De-identified data that underlie the results reported in this article will be made available upon reasonable request.

## Acknowledgements

All authors contributed to conceptual ideas and contributed to methodology. TCC, EPE, JRN contributed to formal analysis; TCC, JS contributed to writing and original draft preparation; All authors contributed to writing, reviewing, and editing the manuscript; JS supervised the work. The authors are grateful to all Kaiser Permanente members without whom this study would not have been possible.

This work was supported by the National Cancer Institute Serological Sciences Network (SeroNet) [U01 CA260584 to JS, LHK, DAC]; The Permanente Medical Group Delivery Science Grant (JS, LHK, DAC), and the Physician Researcher Program of The Permanente Medical Group Delivery Science and Applied Research Program (JS).

All authors: No conflicts of interest identified. All authors have submitted the ICMJE Form for Disclosure of Potential Conflicts of Interest. Conflicts that the editors consider relevant to the content of the manuscript have been disclosed.

---

**Supplemental figure 1.**
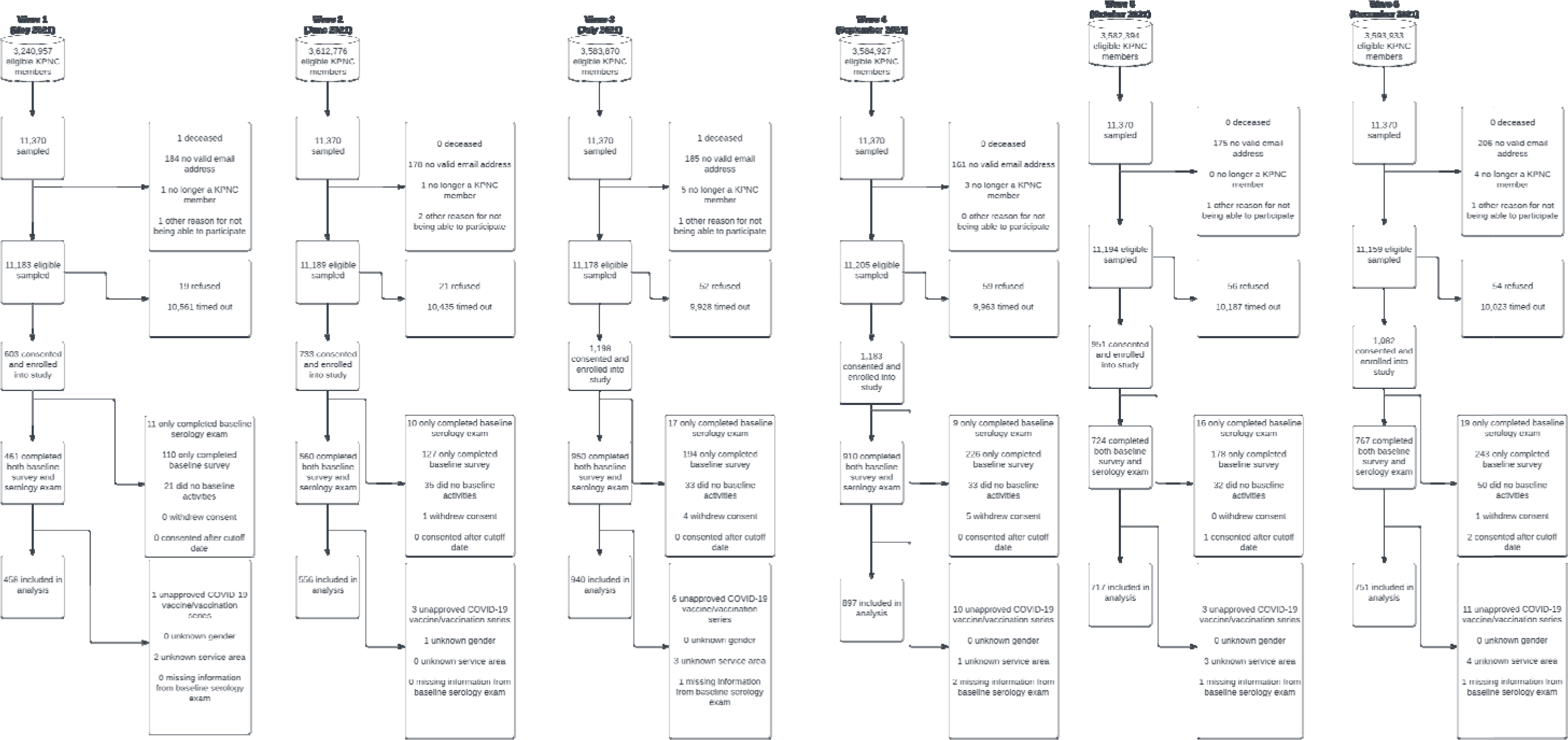
Sampling, inclusions, and exclusions from the Kaiser Permanente Northern California member population May 2021 – December 2021 that lead to the creation of our study population. To be eligible for sampling, members needed to be enrolled in Kaiser Permanente Northern Calfornia membership for at least two months prior to the sampling date, be at least 7 years old on the sampling date, not listed in the “Do not contact” file for study participation, and had not been invited to a previous to a previous Kaiser Permanente Northern California COVID-19 serology study. Wave 1 only included English speaking members.

**Supplementary Table 1:** Characteristics of the eligible Kaiser Permanente Northern California (KPNC) population May 2021 – April 2022 and non-respondent sampled population compared to respondent population before and after weighting.

| Characteristic | Before non-response weighting |  |  | After non-response weighting |  |  | Comparison with KPNC population |  |  |
| --- | --- | --- | --- | --- | --- | --- | --- | --- | --- |
|  | Non-respondent population, N = 63,269 | Respondent population, N = 4,319 | SMD | Non-respondent population, N = 63,269 | Respondent population, N = 4,319 | SMD | Eligible KPNC population, N = 3,498,764 | Respondent population, N = 3,571,275 | SMD |
| **Age group (years)** |  |  | 0.64 |  |  | 0.14 |  |  | 0.10 |
| 7-17 | 6,319 (10%) | 72 (1.7%) |  | 6,319 (10%) | 72 (7.0%) |  | 333,650 (9.5%) | 250,210 (7.0%) |  |
| 18-44 | 27,250 (44%) | 1,062 (25%) |  | 27,250 (44%) | 1,062 (41%) |  | 1,478,525 (42%) | 1,467,581 (41%) |  |
| 45-64 | 18,943 (29%) | 1,743 (40%) |  | 18,943 (29%) | 1,743 (32%) |  | 1,051,877 (30%) | 1,155,306 (32%) |  |
| 65+ | 10,757 (17%) | 1,442 (34%) |  | 10,757 (17%) | 1,442 (20%) |  | 634,712 (18%) | 698,178 (20%) |  |
| **Sex** |  |  | 0.23 |  |  | 0.07 |  |  | 0.06 |
| Female | 33,239 (52%) | 2,751 (63%) |  | 33,239 (52%) | 2,751 (56%) |  | 1,840,164 (53%) | 1,983,715 (56%) |  |
| Male | 30,030 (48%) | 1,568 (37%) |  | 30,030 (48%) | 1,568 (44%) |  | 1,658,599 (47%) | 1,587,559 (44%) |  |
| **Race/Ethnicity** |  |  | 0.65 |  |  | 0.10 |  |  | 0.06 |
| Asian | 11,595 (21%) | 493 (12%) |  | 11,595 (21%) | 493 (21%) |  | 720,161 (21%) | 750,752 (21%) |  |
| Black | 11,899 (7.0%) | 352 (2.8%) |  | 11,899 (7.0%) | 352 (6.0%) |  | 234,344 (6.7%) | 215,024 (6.0%) |  |
| Hispanic | 11,777 (21%) | 371 (8.9%) |  | 11,777 (21%) | 371 (19%) |  | 690,766 (20%) | 662,280 (19%) |  |
| Other | 4,963 (10%) | 156 (4.4%) |  | 4,963 (10%) | 156 (8.9%) |  | 346,479 (9.9%) | 318,404 (8.9%) |  |
| White | 23,035 (41%) | 2,947 (71%) |  | 23,035 (41%) | 2,947 (45%) |  | 1,507,014 (43%) | 1,624,815 (45%) |  |
| **Geographic Area** |  |  | 0.21 |  |  | 0.06 |  |  | 0.05 |
| Central Valley | 7,361 (12%) | 326 (7.2%) |  | 7,361 (12%) | 326 (9.9%) |  | 401,410 (11%) | 355,054 (9.9%) |  |
| East Bay | 18,319 (27%) | 1,169 (27%) |  | 18,319 (27%) | 1,169 (28%) |  | 950,281 (27%) | 1,000,824 (28%) |  |
| North Bay | 3,941 (6.8%) | 458 (11%) |  | 3,941 (6.8%) | 458 (6.9%) |  | 248,204 (7.1%) | 244,676 (6.9%) |  |
| North Valley | 17,485 (27%) | 1,220 (28%) |  | 17,485 (27%) | 1,220 (27%) |  | 936,180 (27%) | 973,315 (27%) |  |
| Peninsula | 7,006 (12%) | 516 (12%) |  | 7,006 (12%) | 516 (12%) |  | 411,419 (12%) | 410,996 (12%) |  |
| South Bay | 9,157 (16%) | 630 (15%) |  | 9,157 (16%) | 630 (16%) |  | 551,270 (16%) | 586,408 (16%) |  |
| **Charlson score category** |  |  | 0.40 |  |  | 0.16 |  |  | 0.14 |
| No visits in prior year | 7,671 (13%) | 125 (3.1%) |  | 7,671 (13%) | 125 (7.8%) |  | 419,085 (12%) | 279,049 (7.8%) |  |
| Score 0 | 39,155 (62%) | 2,647 (62%) |  | 39,155 (62%) | 2,647 (64%) |  | 2,182,429 (62%) | 2,297,304 (64%) |  |
| Score 1 | 8,133 (12%) | 749 (17%) |  | 8,133 (12%) | 749 (13%) |  | 446,424 (13%) | 482,110 (13%) |  |
| Score 2 | 3,276 (5.0%) | 370 (8.4%) |  | 3,276 (5.0%) | 370 (5.6%) |  | 182,307 (5.2%) | 199,918 (5.6%) |  |
| Score 3+ | 5,034 (7.5%) | 428 (9.7%) |  | 5,034 (7.5%) | 428 (8.8%) |  | 268,517 (7.7%) | 312,894 (8.8%) |  |
| **Chronic obstructive pulmonary disease** | 6,514 (9.7%) | 582 (13%) | 0.11 | 6,514 (9.7%) | 582 (11%) | 0.04 | 347,546 (9.9%) | 388,816 (11%) | 0.03 |
| **Chronic kidney disease** | 2,960 (4.4%) | 246 (5.5%) | 0.05 | 2,960 (4.4%) | 246 (4.9%) | 0.03 | 155,094 (4.4%) | 175,862 (4.9%) | 0.02 |
| **Atherosclerotic cardiovascular disease** | 7,195 (11%) | 769 (18%) | 0.19 | 7,195 (11%) | 769 (12%) | 0.02 | 406,301 (12%) | 425,290 (12%) | 0.01 |
| **Cancer/Malignancies** | 1,495 (2.3%) | 196 (4.5%) | 0.12 | 1,495 (2.3%) | 196 (3.3%) | 0.06 | 84,570 (2.4%) | 116,343 (3.3%) | 0.05 |
| **Diabetes** | 6,447 (9.6%) | 441 (9.8%) | 0.00 | 6,447 (9.6%) | 441 (9.6%) | 0.00 | 337,823 (9.7%) | 343,536 (9.6%) | 0.00 |
| **Body mass index category** |  |  | 0.47 |  |  | 0.18 |  |  | 0.15 |
| 18.5 - <25 | 15,581 (26%) | 1,281 (31%) |  | 15,581 (26%) | 1,281 (27%) |  | 910,036 (26%) | 961,658 (27%) |  |
| 25 - <30 | 17,390 (28%) | 1,476 (34%) |  | 17,390 (28%) | 1,476 (30%) |  | 989,570 (28%) | 1,083,245 (30%) |  |
| 30 - <40 | 16,129 (24%) | 1,174 (26%) |  | 16,129 (24%) | 1,174 (27%) |  | 853,304 (24%) | 947,381 (27%) |  |
| >= 40 | 3,858 (5.2%) | 254 (5.4%) |  | 3,858 (5.2%) | 254 (5.3%) |  | 183,351 (5.2%) | 189,048 (5.3%) |  |
| Unknown | 10,311 (17%) | 134 (3.2%) |  | 10,311 (17%) | 134 (11%) |  | 562,502 (16%) | 389,942 (11%) |  |
| **Immunity status at baseline** |  |  | 0.54 |  |  | 0.28 |  |  | 0.25 |
| Neither | 14,601 (22%) | 195 (4.3%) |  | 14,601 (22%) | 195 (13%) |  | 720,428 (21%) | 458,920 (13%) |  |
| Prior infection only | 1,604 (2.4%) | 81 (1.8%) |  | 1,604 (2.4%) | 81 (5.0%) |  | 82,603 (2.4%) | 178,472 (5.0%) |  |
| Vaccinated and prior infection | 3,180 (5.0%) | 326 (7.4%) |  | 3,180 (5.0%) | 326 (7.2%) |  | 182,011 (5.2%) | 257,158 (7.2%) |  |
| Vaccinated only | 43,884 (71%) | 3,717 (86%) |  | 43,884 (71%) | 3,717 (75%) |  | 2,513,721 (72%) | 2,676,725 (75%) |  |
| **Vaccination series at baseline** |  |  | 0.62 |  |  | 0.16 |  |  | 0.12 |
| Not vaccinated | 16,247 (24%) | 289 (6.4%) |  | 16,247 (24%) | 289 (19%) |  | 806,138 (23%) | 667,483 (19%) |  |
| Partial | 2,504 (3.8%) | 43 (0.9%) |  | 2,504 (3.8%) | 43 (2.8%) |  | 125,733 (3.6%) | 100,174 (2.8%) |  |
| Primary series complete | 39,680 (64%) | 3,162 (73%) |  | 39,680 (64%) | 3,162 (69%) |  | 2,259,042 (65%) | 2,454,131 (69%) |  |
| Primary series + additional dose | 4,838 (8.0%) | 825 (19%) |  | 4,838 (8.0%) | 825 (9.8%) |  | 307,851 (8.8%) | 349,486 (9.8%) |  |
| **Vaccine type at baseline** |  |  | 0.54 |  |  | 0.16 |  |  | 0.13 |
| Ad26.COV2.S (Janssen) | 3,000 (4.7%) | 192 (4.4%) |  | 3,000 (4.7%) | 192 (4.2%) |  | 164,981 (4.7%) | 148,658 (4.2%) |  |
| BNT162b2 (Pfizer/BioNTech) | 27,790 (45%) | 2,107 (49%) |  | 27,790 (45%) | 2,107 (45%) |  | 1,578,260 (45%) | 1,610,704 (45%) |  |
| mRNA-1273 (Moderna/NIH) | 16,232 (26%) | 1,731 (40%) |  | 16,232 (26%) | 1,731 (32%) |  | 949,384 (27%) | 1,144,430 (32%) |  |
| Unvaccinated | 16,247 (24%) | 289 (6.4%) |  | 16,247 (24%) | 289 (19%) |  | 806,138 (23%) | 667,483 (19%) |  |

KPNC = Kaiser Permanente Northern California; SMD = standardized mean difference
Sampling weights were calculated as the inverse of the probability of selection in a given age and race strata. We adjusted for non-response bias by calculating a response weight defined as the inverse of the probability of having completed the baseline serology exam and survey. This probability was calculated using the “SuperLearner” package for the R programming language. SuperLearner evaluates several candidate algorithms for model fitting and creates a weighted ensemble model from these candidate algorithms. The algorithms considered in our weighting process were: lasso regression, logistic regression, Pearson’s correlation coefficient, generalized additive models, random forest, gradient boosted decision trees, and a simple mean as a baseline. Each algorithm was fitted with the complete covariate set as well as reduced covariate sets determined by screeners for relationships with the outcome, specifically the univariate Pearson correlation, LASSO regression, and random forest variable importance measures.
The variables used in non-response weighting were race/ethnicity, gender, KPNC service region, age group, Charlson comorbidity score category, binary ever received a COVID-19 vaccine, recruitment wave, body mass index category, baseline immunity status, and multi-level vaccination status. Non-response weights were normalized to the size of the eligible sampled population and trimmed such that the maximum trimmed weight was the 99th percentile of the untrimmed weights. The final weight used in the analysis was the product of the sampling weight and the non-response weight.

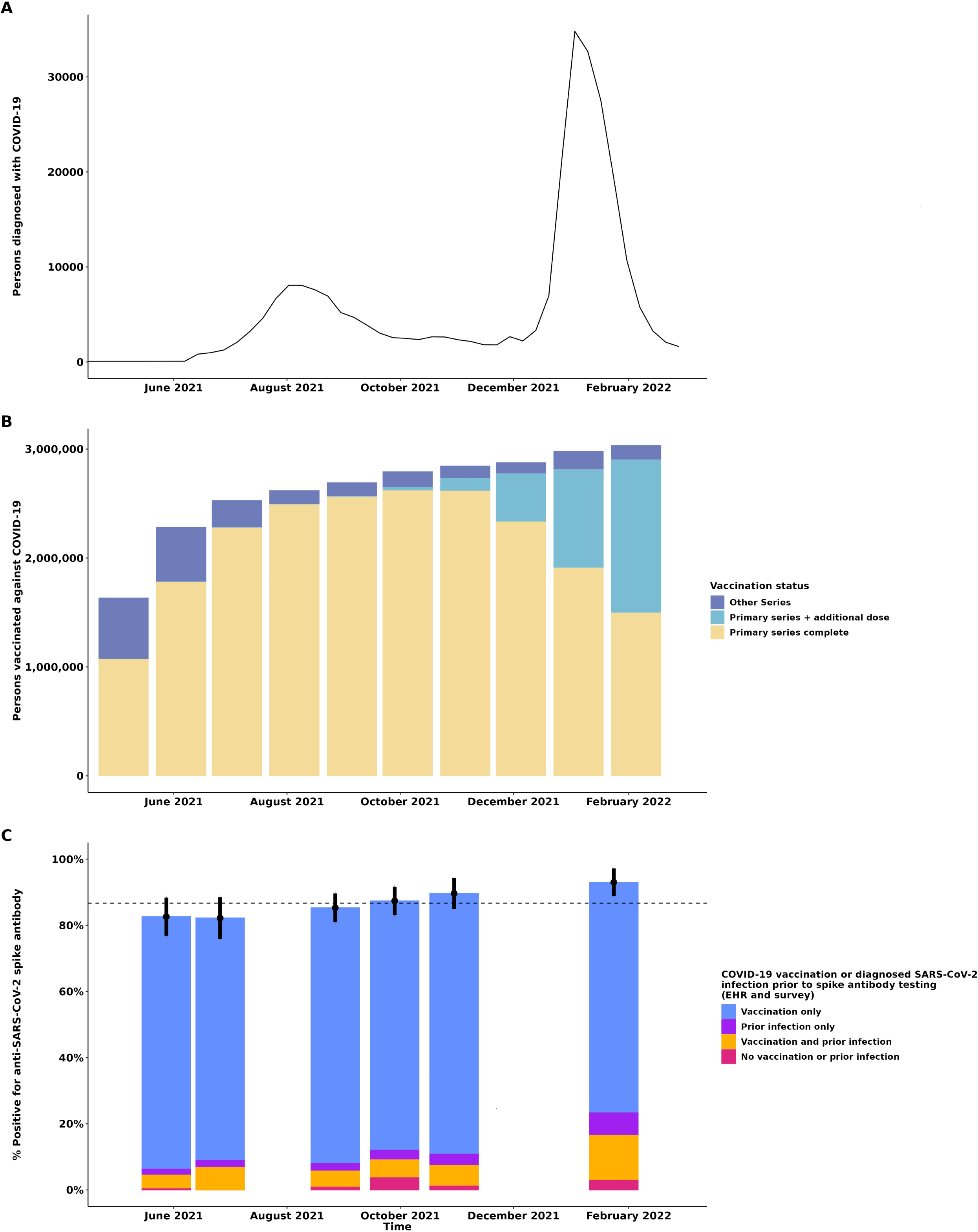

