## Supplementary figures and images for "Relative contribution of COVID-19 vaccination and SARS-CoV-2 infection to population-level seroprevalence of SARS-CoV-2 spike antibodies in a large integrated health system"

### Supplemental table

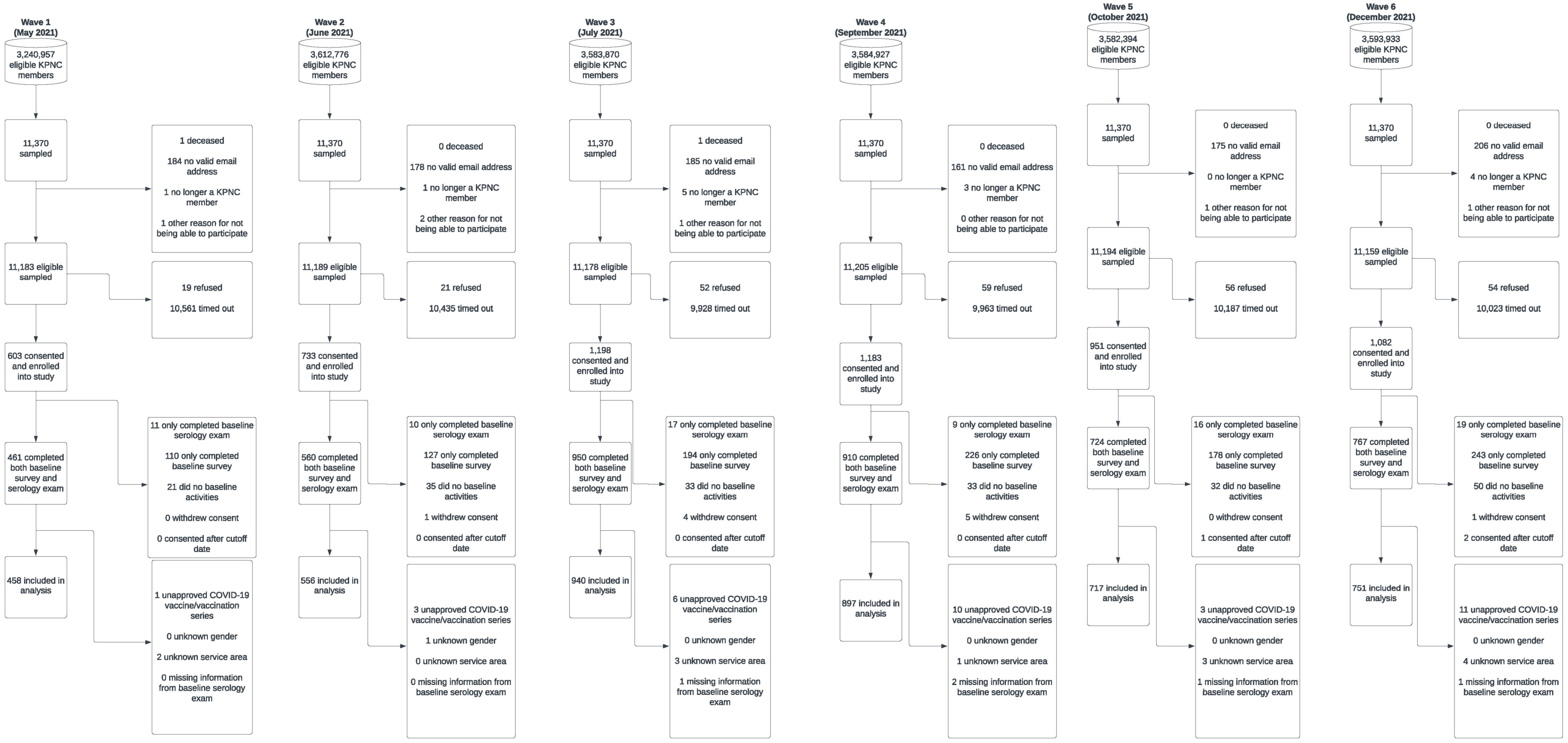
